# Performance of Handcrafted Radiomics versus Deep Learning for Prognosticating Head and Neck Squamous Cell Carcinoma – A Systematic Review with Critical Appraisal of Quantitative Imaging Studies

**DOI:** 10.1101/2024.10.22.24315007

**Authors:** Varsha Gouthamchand, Louise AF Fonseca, Frank JP Hoebers, Rianne Fijten, Andre Dekker, Leonard Wee, Hannah Mary Thomas T

## Abstract

Head and neck squamous cell carcinoma (HNSCC) presents a complex clinical challenge due to its heterogeneous nature and diverse treatment responses. This systematic review critically appraises the performance of handcrafted radiomics (HC) and deep learning (DL) models in prognosticating outcomes in HNSCC patients treated with (chemo)-radiotherapy. A comprehensive literature search was conducted up to May 2023, identifying 23 eligible studies that met the inclusion criteria of methodological rigor and long-term outcome reporting. The review highlights the methodological variability and performance metrics of HC and DL models in predicting overall survival (OS), loco-regional recurrence (LRR) and distant metastasis (DM). While DL models demonstrated slightly superior performance metrics compared to HC models, the highest methodological quality was observed predominantly in studies using HC radiomics. The findings underscore the necessity for methodological improvements, including pre-registration of protocols and assessment of clinical utility, to enhance the reliability and applicability of radiomic-based prognostic models in clinical practice.

## Introduction

Head and neck squamous cell carcinomas (HNSCC) comprise a highly heterogeneous subset of neoplastic diseases originating in the mucosal lining of the oral cavity, pharynx, and larynx [1]. According to GLOBOCAN 2022, HNSCC (including cancers of the lip, oral cavity, larynx, oropharynx, hypopharynx, and salivary glands) accounted for an estimated 826,040 new cases and 445,896 deaths, representing about 4.4% of all cancer cases and 4.6% of all cancer deaths globally [2]. The growing global health burden may be due to alcohol and nicotine consumption patterns correlated with urban/rural migration, economic factors influencing changes in dietary patterns) and a wider exposure to oncogenic viruses such as the human papilloma virus (HPV) in the case of oropharyngeal carcinoma.

Locally advanced HNSCC are generally treated with a combination of radiotherapy (RT), chemotherapy and/or surgery. Prognoses for 5-year survival range from almost 90% in HPV-positive oropharyngeal carcinoma (OPC) down to 25% in advanced hypopharyngeal carcinoma (HPC). Long-term side-effects of treatment also vary considerably between persons and may include physical appearance changes (mainly due to surgery), xerostomia, dysphagia, odynophagia, fibrosis, fatigue, and ototoxicity (mainly due to cisplatin chemotherapy). Survivorship within certain subtypes of HNSCC has been improving gradually over time, leading to greater attention towards functional preservation after treatment, psychosocial resilience, and health-related quality of life. Newer treatments such as immunotherapy and proton beam therapy are not readily available in all countries, therefore great diligence is required to identify patients that benefit from expensive novel treatments, or else to reduce disutility of care among patients that do not benefit from aggressive treatment.

Genetic diversity and complex pathophysiology imply significant intra- and inter-tumoral heterogeneity in HNSCC (add REF here). Routine oncological imaging with computed tomography (CT), positron emission tomography (PET) and magnetic resonance imaging (MRI) are broadly limited to qualitative (visual) interpretation of the images and/or highly simplified metrics (e.g. measuring the maximum tumor diameter on a single axial slice or using the maximum tracer uptake intensity). The added value of clinical imaging in cancer management is unquestionable, but it remains unclear if such non-invasive investigation sufficiently captures the complicated phenotype of HNSCC to guide risk-based stratification.

Radiomics has emerged as a prominent tool for scientific investigation and prognostic modelling of cancer outcomes. Radiomics uses large numbers of quantitative features per subject extracted by a computer algorithm from annotated regions of interest (ROIs) in CT, MRI and PET images [3–5]. More recently, deep learning neural networks (DLNNs) [6, 7] have delivered many significant advances in the field of computerized image analysis, hence DLNN-based oncology outcome modelling is now a rapidly growing research topic. The former requires pre-defined mathematical functions to be evaluated on a region of interest in the image, the so-called “hand-crafted (HC) features” approach. In contrast, DLNNs abstract image information as “deep-learning (DL) features” through a consecutive sequence of local convolution and max-pooling steps. Thus, the latter is considered an exclusively data-driven or knowledge-agnostic approach that does not require pre-definition of features.

From 2020 onwards, there have been many reviews about HC radiomics and DL for HNSCC prognostication, indicating growing interest and rapid innovations in this field of study [8–16]. Giraud et al. [17] conducted a wide-ranging overview of machine learning for radiotherapy applications (including radiomics) in head and neck cancers but did not provide a systematized synthesis of evidence nor detailed critical appraisals of methodological quality. Spadarella et al. [18] supplied a systematic review of radiomics for nasopharyngeal carcinoma for MRI only. Sanduleanu et al. [19] proposed a radiomics quality scoring system however calibration of this scoring scale remains uncertain. Other authors pointed out possible problems of reducing something as highly nuanced and complex as study quality into a single value [20, 21]. Guha et al. [22] systematically reviewed the radiomics literature up till February 2018 for effectiveness of treatment but did not explicitly search for DL imaging studies. Despite these valuable contributions, a critically appraised synthesis comparing both radiomics and deep learning for HNSCC prognostication, covering a broad range of imaging modalities and addressing methodological rigor, remains lacking.

The central question addressed in this systematic review with critical appraisal of methodological quality is to estimate the ***discriminative performance envelopes of prognostic HNSCC models employing handcrafted radiomics (HC) and/or deep learning (DL)***. The performance data shall be gleaned from high-quality primary research articles containing long-term treatment outcomes in locally advanced primary OPC, HPC and laryngeal carcinoma (LC) that are widely treated by (chemo-) radiotherapy, either alone or post-operatively. The primary result expected is a body of evidence for relative efficacy of HC versus DL features for the prognostication tasks in HNSCC

In this review we do not cover: (i) studies that are principally about nasopharyngeal carcinoma because it is an epidemiologically distinct disease, and (ii) local cancers in the oral cavity that are managed with surgery alone. We placed emphasis on the methodological reliability of each study and summarized the reviewed models.

## Methods

A protocol for this systematic review has not been prospectively registered on a database before performing it.

### Eligibility criteria

Eligible studies include only human subjects diagnosed with primary HNSCC that have been treated by (chemo)-radiotherapy either alone or in combination with surgery and post-operative RT. This clinical setting was selected because of nominally standardized and quality-assured protocols (particularly of radiotherapy planning CT that are needed for radiation dosimetry calculations) along with expertly outlined Gross Tumor Volume (GTV) as the region of interest (ROI) by the practicing clinicians.

Studies must report at least one clinical outcome (such as all-causes mortality, cancer-related mortality, progression, regression, local and/or regional failure, or distant metastasis). Articles eligible for review contained: (i) HC radiomics and/or DL features derived from pre-treatment clinical imaging, and (ii) TRIPOD 3B (development and validation using separate data) or higher (Type 4: validation only) type of investigation of clinical outcomes modelling [23].

### Exclusion criteria

Specific exclusion criteria were: (i) nasopharyngeal carcinoma, (ii) exclusively phantom, *in vitro* or *in silico* studies, and (iii) clinical oncology imaging modality other than CT, PET or MRI.

Studies concerning exclusively short-term response immediately following treatment (such as RECIST criteria, tumor expansion/shrinkage) or studies pertaining exclusively to radiomics/DLNN-based diagnostic characterization (such as epidermal growth factor or HPV expression), but without long-term clinical outcomes, were excluded from the review.

Excluded studies also lacked peer-reviewed full text from the publishing journal, or if published before 1^st^ January 2011, or if full text was not available in the English language.

### Information sources

The primary search for eligible studies up to the end of May 2023 was conducted within the PubMed electronic database after it had been merged with EMBASE. The secondary search was conducted by scanning for eligible studies within the bibliography of reviews and systematic reviews. “Grey” literature sources were not consulted for this review. By-hand searching of individual journal catalogues was not performed. Non-peer reviewed article collections (e.g. arXiV and medArXiV) were omitted from this search.

### Search strategy

For PubMed, a sensitive search for diagnostic and prognostic studies was performed using a combination of the broad Haynes [24] and Ingui [25] filters, with an additional modification proposed by Geersing [26]. The search was narrowed using MeSH term for “head and neck cancer”, or text words anywhere in the title and abstract referring to radiomics and deep learning (including common synonyms). Text word searches were first combined with ‘OR’ operators, then integrated to MeSH term and prognostics studies filter using ‘AND’ operators. The plain text of our search string is provided as Supplemental Text Box 1. The search was conducted in two phases, in August 2020 and again on March 31^st^, 2021, and all returned records were merged prior to screening. We also checked the references of all the review articles on ‘head and neck prognostication’ for any additional articles that may have been missed in our electronic database search.

### Study selection

We approximated the methodological conventions established by the Cochrane Collaboration for systematic reviews due to the small size of our review team. Two reviewers (VG and LAAF) during the first search phase and two reviewers (VG and HMTT) during the second search phase independently screened PubMed records only by title and abstract to identify potential articles. Disagreements during screening were resolved by unanimous consensus through re-appraisal together with a third reviewer (LW). The full text for candidate articles was obtained through the authors’ institutional library subscriptions. Three reviewers working separately (VG, LW and HMTT) subjected full texts to a detailed reading against inclusion and exclusion criteria, then additional disagreements were resolved by unanimous consensus through reappraisal.

### Data extraction

First, general details of the eligible studies were summarized as tables. These included the primary cancer type, imaging modality and image acquisition details, cohort clinical information with sample size, primary outcomes including non-radiomics and non-deep-learning-based comparator factors, and the software base for HC or DL.

### Estimating risk of bias in individual studies

There have been several tools proposed to appraise methodological quality of prognostic and diagnostic studies in general, such as QUADAS [27], or radiomics-specific score (RQS) [19].

We have based a methodological appraisal on the rationale raised by the RQS but refrained from assigning a single quality number. In its place, we included a brief overview of what, in our view, might have constituted some part of methodological robustness in the study. Each of the three reviewers worked independently on extracting the methodological information and was afterwards cross-checked by another reviewer. The methodological aspects we sought to extract from the studies were the following:

1. Was the study prospectively registered for the intended analysis methodology in a publicly accessible study database prior to commencement of the analysis?
2. If imaging data were not publicly available for download, were sufficient details present in the article to identify the scope of validity (e.g., diversity of equipment vendors, whether i.v. contrast was used, image acquisition parameters etc.)?
3. If digital image pre-processing had been applied (digital filters, isotropic resampling, augmentations such as flips or rotations, and related operations) was enough information provided or standardized steps documented, that would support reproducing the same steps independently?
4. If some form of model simplification had been performed, such as feature dimensionality reduction or drop-outs, was enough information provided or standardized steps documented, that would support reproducing the same model simplifications independently?

- Was some form of model interpretability incorporated into the findings, such as biological correlates of HC features, or attention maps of DL features, that could support a high degree of clinical verification of the output?
5. If risk stratification groups had been defined (e.g. cut-offs and operating point) was a clinical justification provided rather than solely relying on model fine-tuning for optimal groupings, since the latter might produce overly optimistic results of discrimination?
6. Was the reference standard of outcomes used in the supervised learning (also known as ground truth) provided by human experts <u>and</u> closely matched with the context of the clinical decision being supported by the proposed model?
7. Whether the expected clinical utility of the proposed model had been estimated, through some form of cost-benefit or decision-curve analysis or related measure of decision-making utility?

## Results

### Study selection

The PRISMA (Preferred Reporting Items for Systematic review and Meta-Analysis) [28, 29] flowchart is provided as Figure 1. 1610 records were identified based on the specified search terms in PubMed and 237 additional records through other sources. After duplicates removal, there were 1718 articles available for screening. Applying the selection criteria led to 120 studies for full-text screening. In the end, 23 articles were deemed eligible.

**Fig 1.**
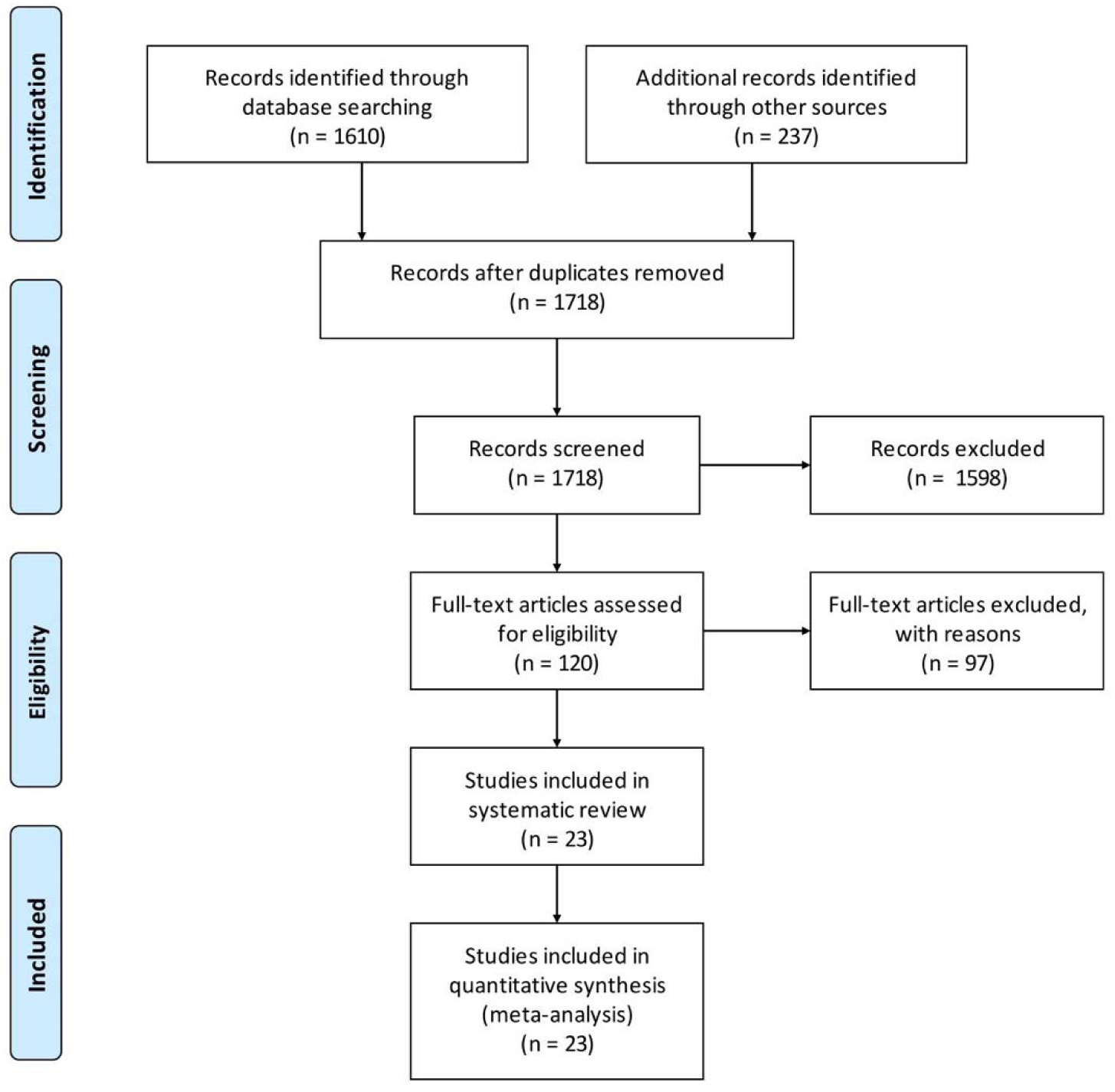
Meta-analyses (PRISMA) workflow to select articles for review.

### Characteristics of included studies

Table 1 presents an overview of the general characteristics observed in all the studies included. The majority (16/23) of the studies included HC radiomics, six studies included DL, and one study involved both HC and DL features for prediction of outcome following radiation therapy for HNSCC.

**Table 1.**
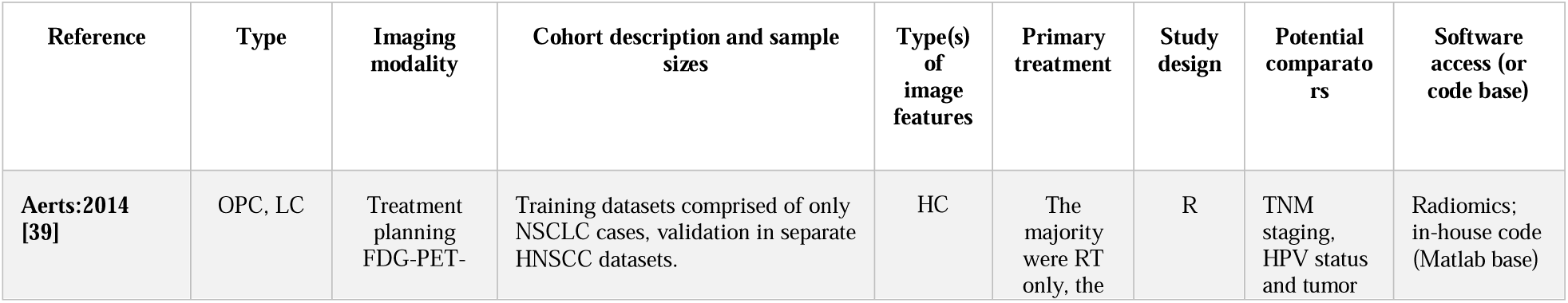
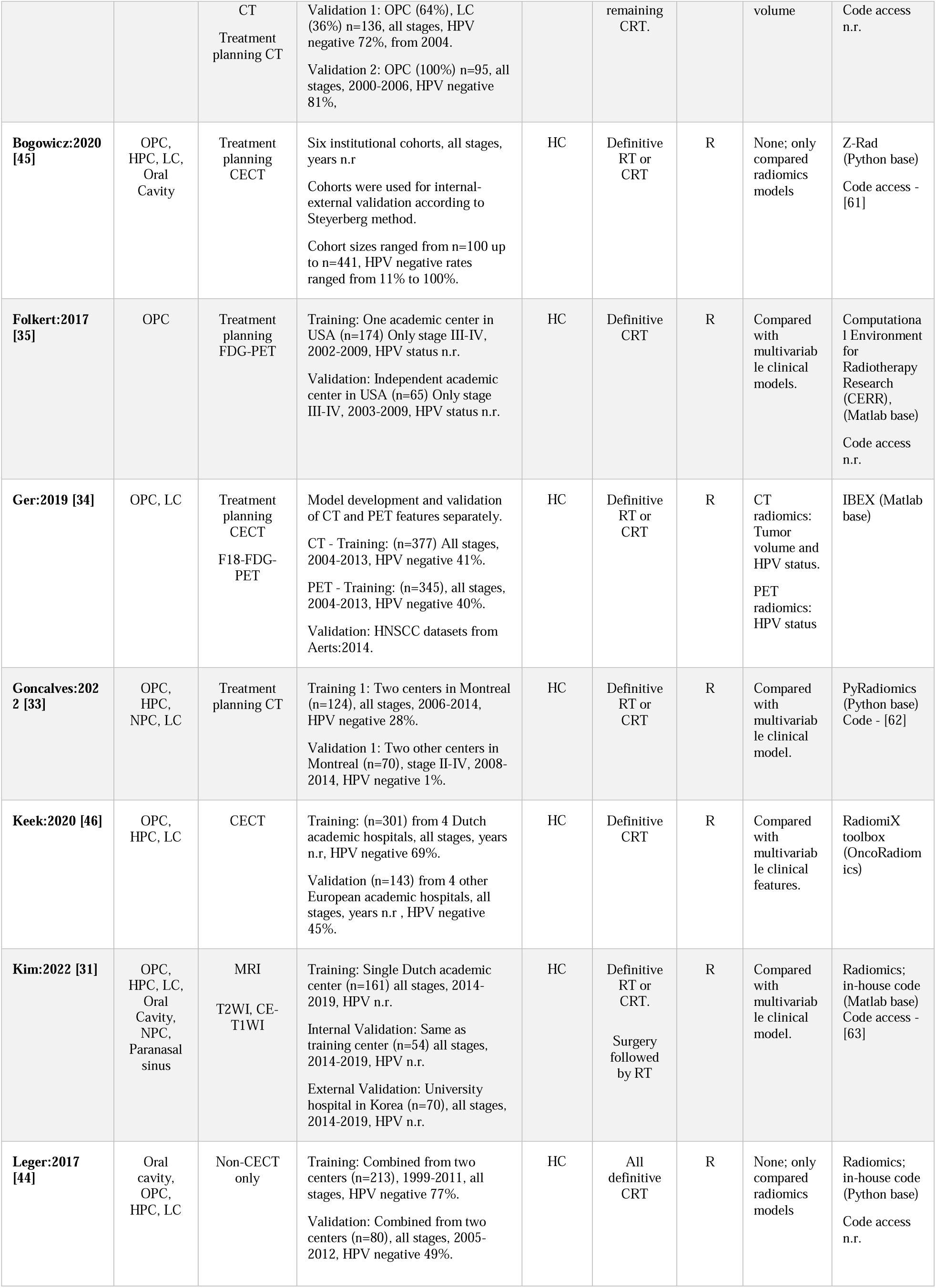
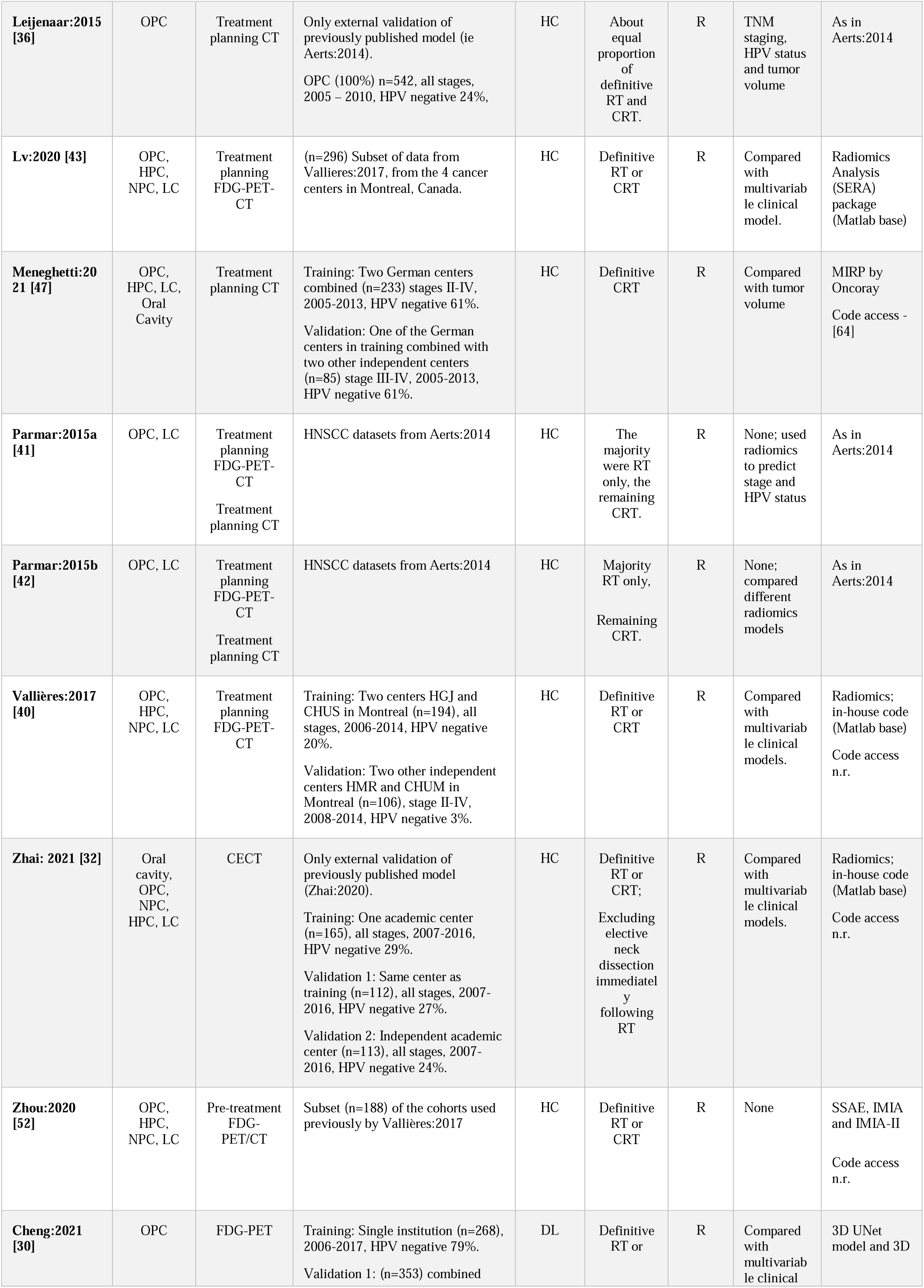
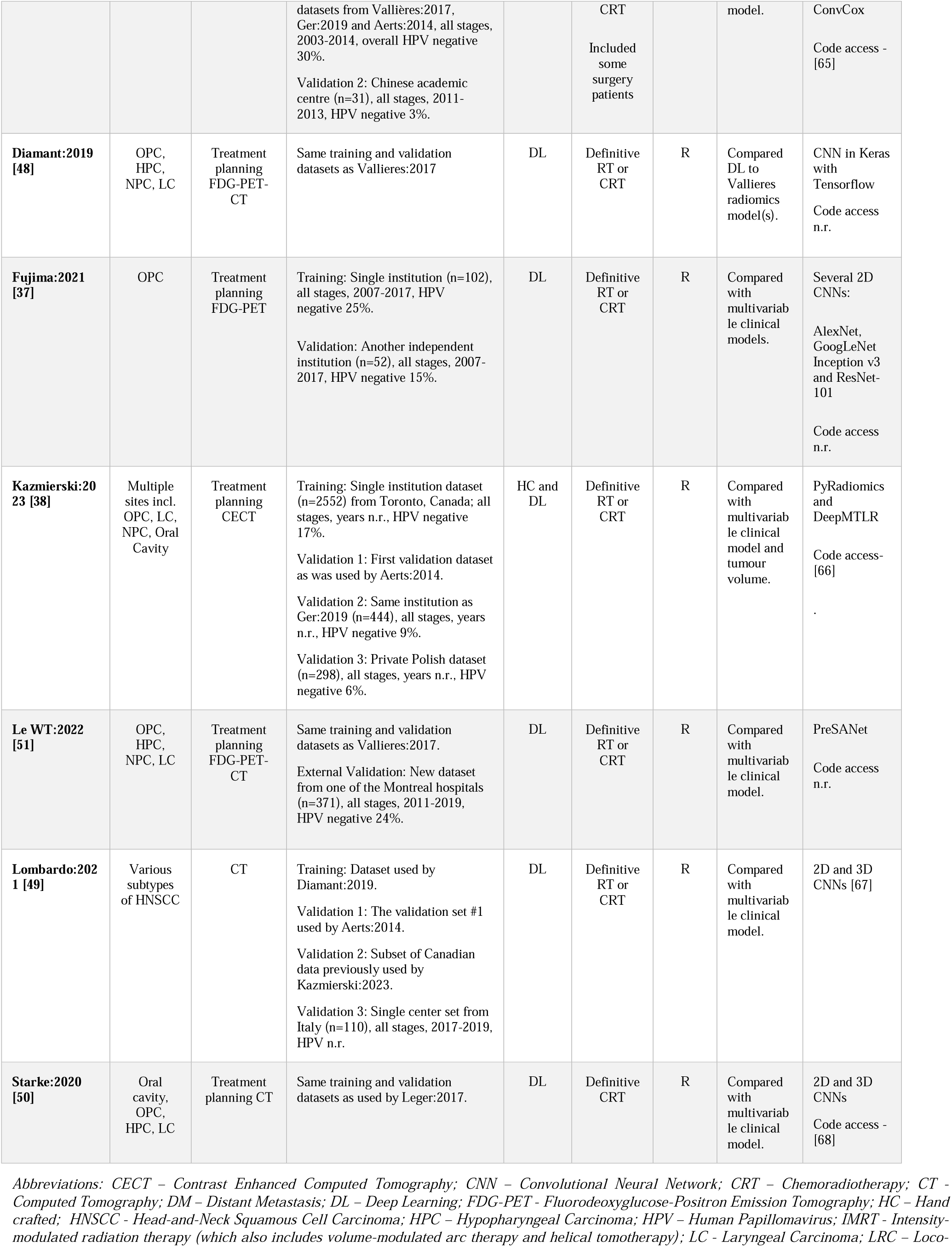
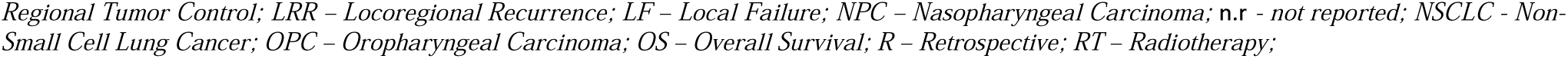
Summary of general study characteristics.

The most widely reported disease subsite was oropharyngeal cancer with almost all studies including this in their dataset (22/23). Next major tumor site represented was larynx (18/23), followed by hypopharynx (13/23) and oral cavity (7/23). Although we did not specifically include nasopharyngeal cancer, some studies reported a mixed subset of patients with NPC (9/23) and paranasal cancer (1/23) in the training or validation cohorts.

Most of the studies included radiomics derived from radiotherapy treatment planning images; some of them included FDG PET-CT (8/23), FDG PET (4/23) or CT (14/23). Additionally, only one study reported the use of MRI. Only 5 studies reported the use of contrast-enhanced CT images.

Most of the patients included in the studies were treated with definitive radiotherapy or chemo-radiation therapy. Cheng et al. included some patients who underwent surgery [30]. One study included patients who had surgery following RT [31]. Zhai et al. specifically excluded patients who underwent elective neck node dissection following RT [32].

The sample size of the cohorts reported in this review ranged from 52 to 2552. For HC radiomics studies used training dataset sizes ranging from 124 [33] to 377 [34], (mean 202; SD 101), and their validation dataset sizes varied from 65 [35] to 542 [36], (Mean 143; SD 107). In the DL studies, the training datasets sizes ranged from 102 [37] to 2552 patients [38], (Mean 531; SD 826) and independent validation datasets from 52 [37] to 872 patients [38], (Mean 200; SD 133).

A wide variety of software tools were used to extract the HC features, with 9 studies reporting the use of custom-built codes using MATLAB [31, 32, 36, 39–43] or Python [44]. The other open-source software reported were Z-Rad [45], IBEX [34], CERR [35], OncoRadiomics [46], Pyradiomics [33] and MIRP [47].

Most DL studies [30, 37, 48–50] applied a reasonably consistent CNN architecture, from what could be gleaned in the technical details of the publications. For instance, Le at al. used a 3-layer neural network with self-attention, also known as PreSANet (Pre-Self-Attention Network) [51] while Kazmierski et al. used a deep multitask logistic regression to model the time-to-event [38].

### Summarized performances of included studies

Table 2 summarizes the endpoints, model building aspects, and performance of the different models. The most studied prognosis endpoint (16/23) was overall survival (OS) followed by local disease failure, recurrence or control and finally distant metastasis. The event to sample ratios for OS ranged from 15% to 81% and distant metastasis (DM) rates varied between 12% and 19%. The rates for loco-regional recurrence (LRR), local recurrence (LR) local failure (LF) or local regional control (LRC), ranged from 7% to 67%.

**Table 2.**
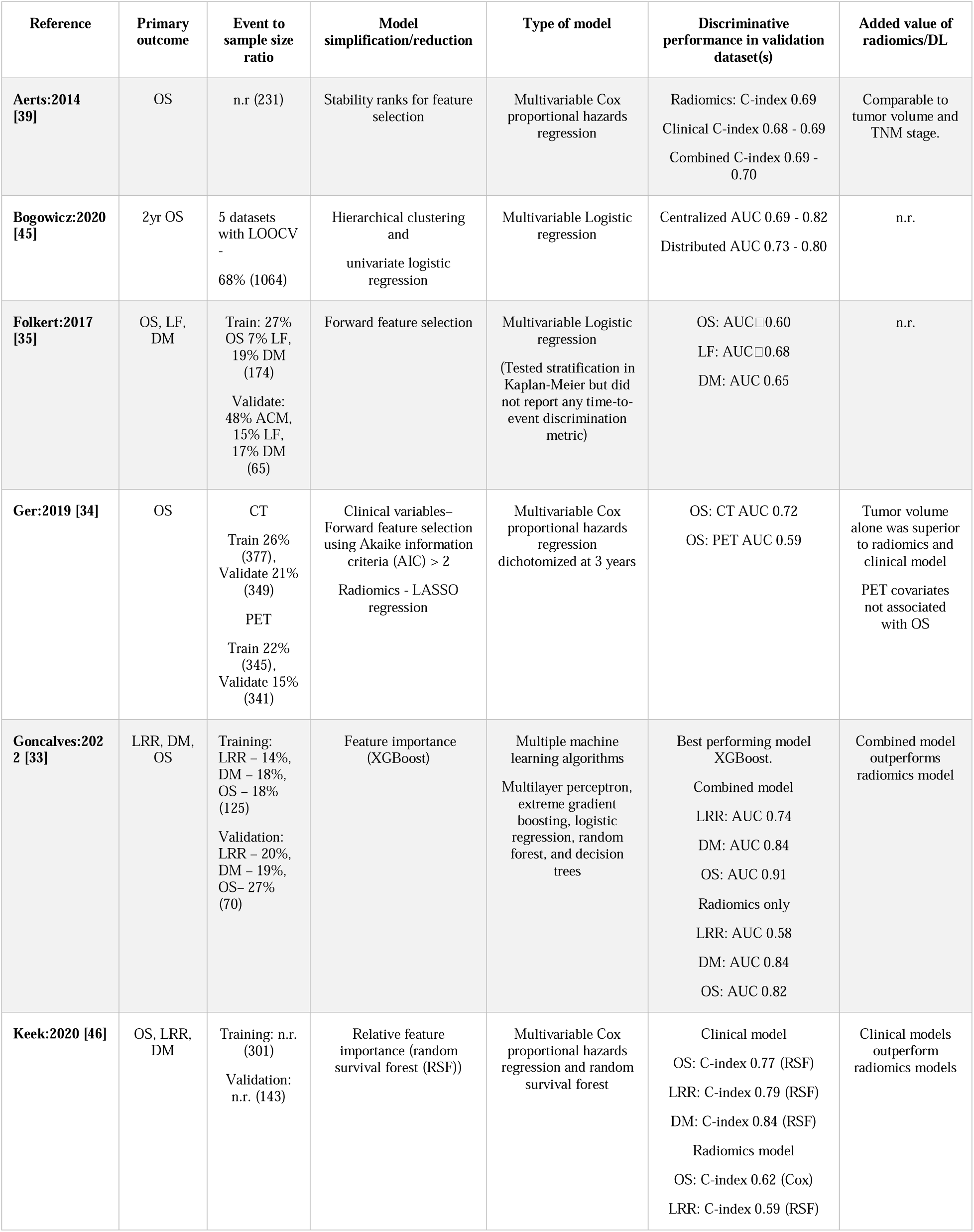
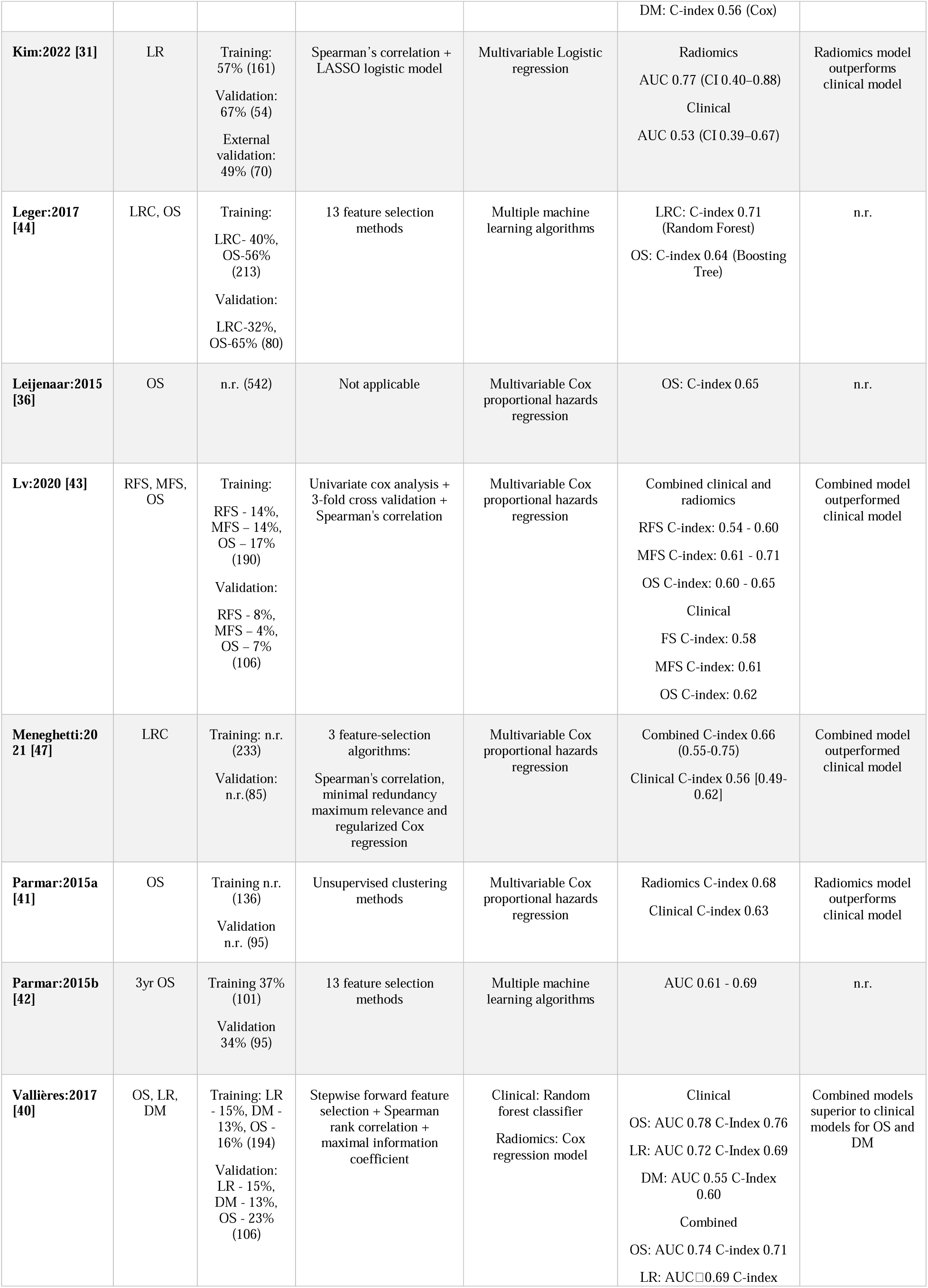
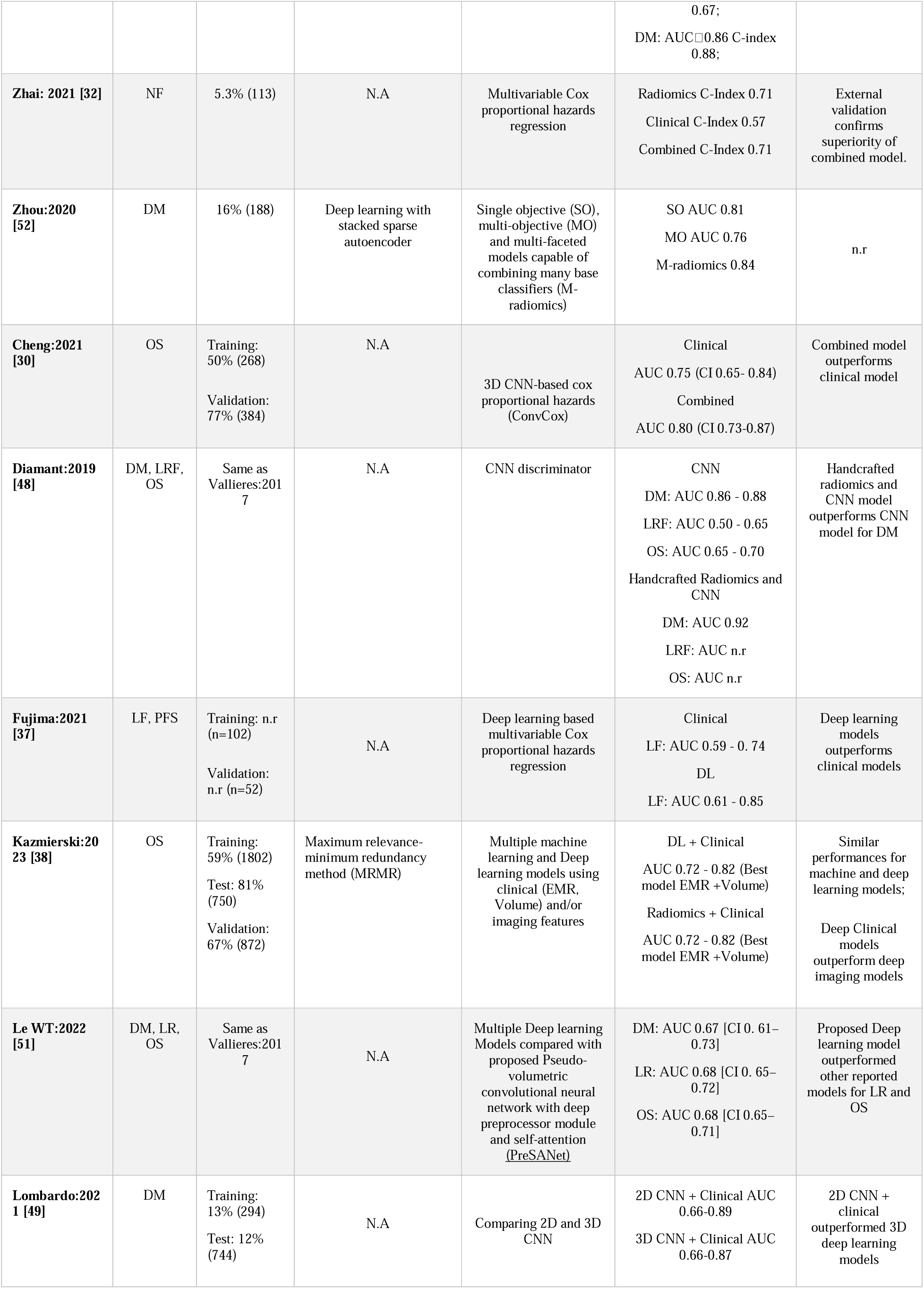
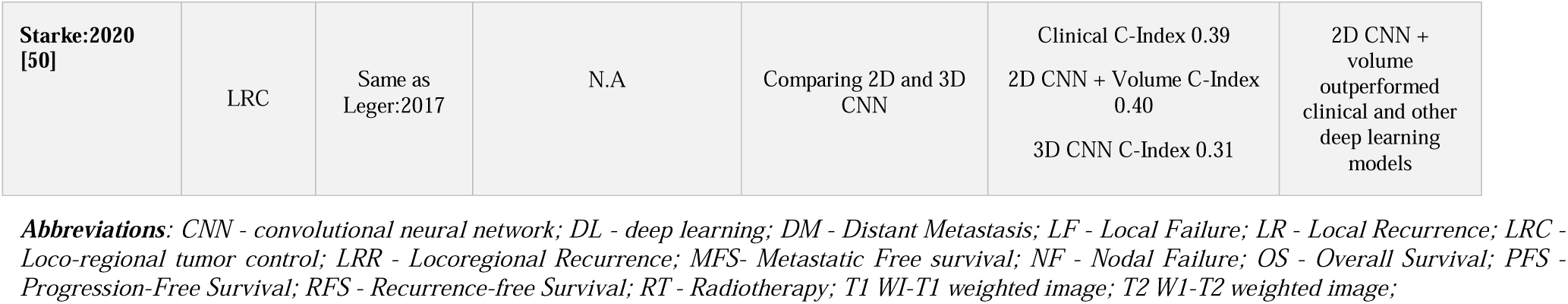
Summary of model discriminative performances.

Notably, all sixteen HC radiomics studies employed various feature reduction techniques to streamline their radiomics models. Among these, Spearman’s correlation ranking with the other features emerged as one of the popular methods [31, 40, 43, 47] followed by LASSO regression models [31, 34]. Some studies employed more than one feature reduction approach before building the prognostic models. For example, [42, 44] utilized 13 feature selection methods for their various machine learning algorithms.

The most frequently (9/23) used machine learning model for HC radiomics was multiple Cox regression technique followed by the multiple linear regression. When we analyzed the prognostic performance of these models across the studies based on reported test AUC/C-Indices, the best performing model for OS was reported by Goncalves et al. with an AUC/C-index of 0.91 using HC features [33]. Analyzing the DL studies separately showed that Kazmierski et al. [38] achieved the highest performing OS model with a discriminatory AUC of 0.82. Vallieres et al. [40] reported the highest performing Distant metastasis (DM) prediction model using HC radiomics and clinical parameters with a C-Index of 0.88 while Lombardo (2021) [49] achieved an AUC of 0.89 by including DL and clinical features. Interestingly, [48] constructed a CNN model using HC radiomics features, which surpassed all other performances and achieved an AUC/C-index of 0.92.

Consequently, the majority of HC studies (7 out of 16) and DL studies (5 out of 6) found that incorporating radiomics, either alone or combined with clinical parameters enhanced the predictive power and offered value compared to the traditional clinical models.

### Methodological quality assessment

Given the large number of HC or DL studies for prognostication in head and neck cancer, we restricted the assessment of methodological quality of studies that adhered to TRIPOD guidelines and described the development or validation of the model or both. (TRIPOD 3B and 4). Figure 2 gives an overview of the distribution of the methodological quality and reporting completeness for 23 studies selected for this review. An extended explanation of the reasons for the scores is made available as part of the supplementary Table S1.

**Fig 2.**
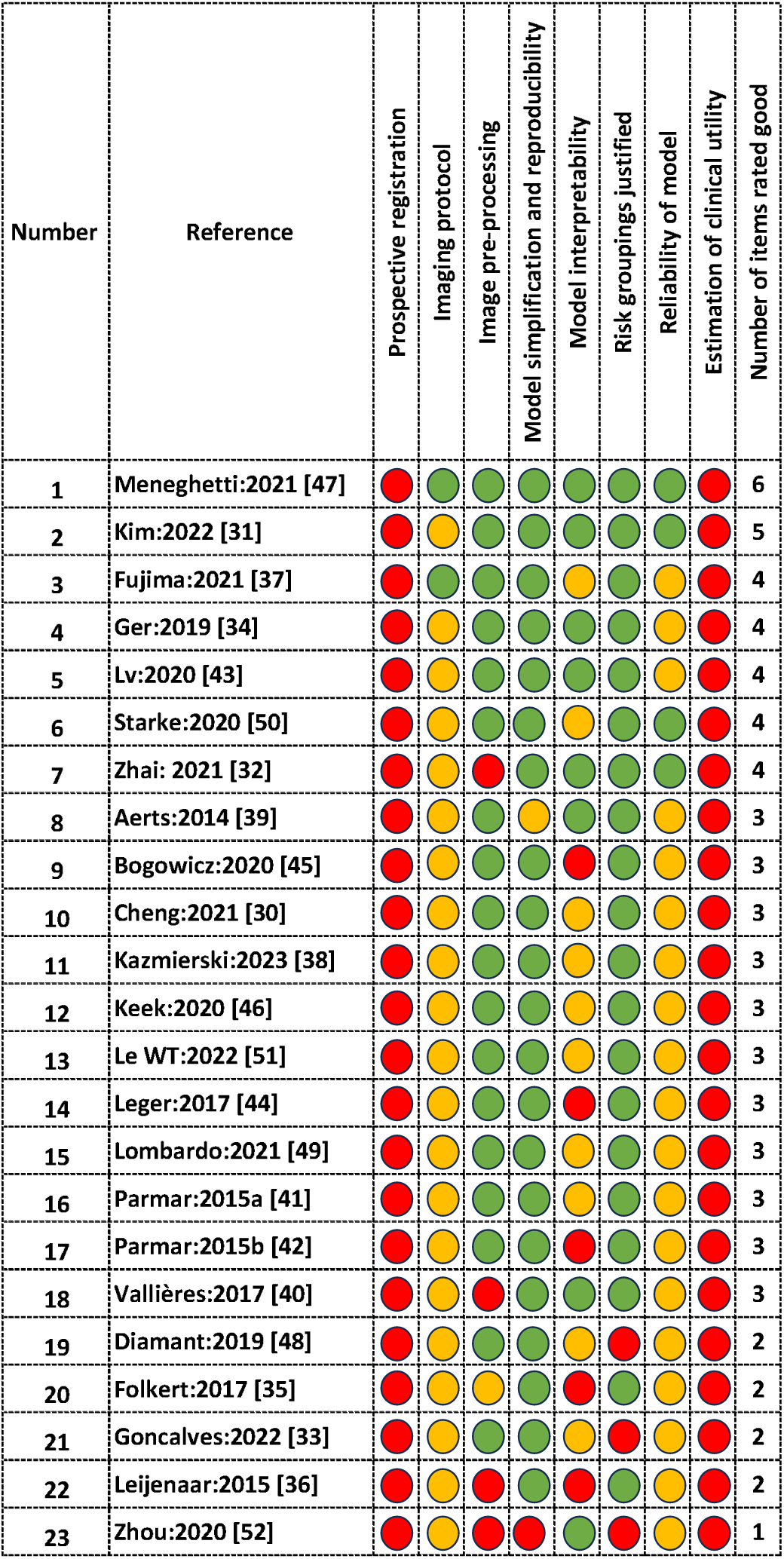
Methodological quality assessment of included studies. Red, yellow, and green dots represent poor, medium, and good quality, respectively. The final column shows the total number of good scores (green) out of 8 quality assessments.

The methodological assessment involved using an 8-item rating system rating based on the criteria as mentioned earlier under the Methods section, with red, yellow, and green indicating poor, medium, and good respectively. The highest score achieved was 6/8 by only one study, falling short due to study not prospectively registered and show the clinical utility of the models [47]. More than half of the reporting was of potentially suboptimal methodological quality and achieved a score of 3 or lower. None of the studies were prospectively registered prior to the HC or DL analysis, which recapitulates a general limitation seen globally in prediction modelling studies using HC features and DL. Only 2/23 studies reported all essential details regarding the imaging acquisition protocol, and 18/23 about any image pre-processing. Regarding assessing the clinical utility of the developed models through methodologies like cost-benefit analysis or decision curve analysis; none of the studies included in this review fulfilled this criterion. Most studies (20/23) included the essential model simplification techniques such as feature selection/multi-dimensionality reduction, hyper-parameters, and dropout rates etc. that allow reproducibility of the models. However, fewer (7/23) studies included measures for interpretability such as comparison with biological correlates. Most studies (20/23) provided appropriate justification for risk grouping/risk cut-offs to delineate risk subgroups; however, three studies [33, 48, 52] did not include any risk stratification.

We found only 4/23 of the studies investigated the overall effectiveness of the models in comparison to the traditional clinical models within the healthcare setting. The broad parameters assessed included were if a) the AI models were compared to clinical models b) the models were trained and tested for the outcomes appropriately c) if the outcome was survival, the survival data used were linked to any cancer registries, and d) adequate documentation of the final model that permits reproducibility for external validation. The two studies that reported ‘good’ in at least 5 out of the 8 assessment items [31, 47] used HC radiomics and were both reporting for local disease as the outcome. Of the five studies that had ratings of 4, 3 used HC radiomics and 2 used DL, all reporting varied outcomes. Most of the other studies (16/23) had ratings of 3 or less, which included 10 studies using HC radiomics, 5 using DL methods and 1 study having both HC and DL features.

Figure 3 visualizes the reported discriminatory metrics (AUC/C-index) against the number of methodological items rated ‘good’ in this review. The color-codes refer to the type of features used for modelling the outcomes, namely HC or DL. The top two methodological rated studies had a discriminatory performance between 0.66-0.77. We noticed that most of the studies lie within a wide scatter with respect to the performances ranging from 0.58-0.92 and ratings between 2 and 4. From three HC radiomics studies, we observed an AUC/C Index of 0.83 to 0.91 [33, 40, 52] for four outcomes. Five DL studies had discriminatory performance between 0.80-0.92 [30, 37, 38, 48, 49]. To view the reported metrics against our ratings for each outcome individually, please refer to the supplementary materials.

**Fig 3.**
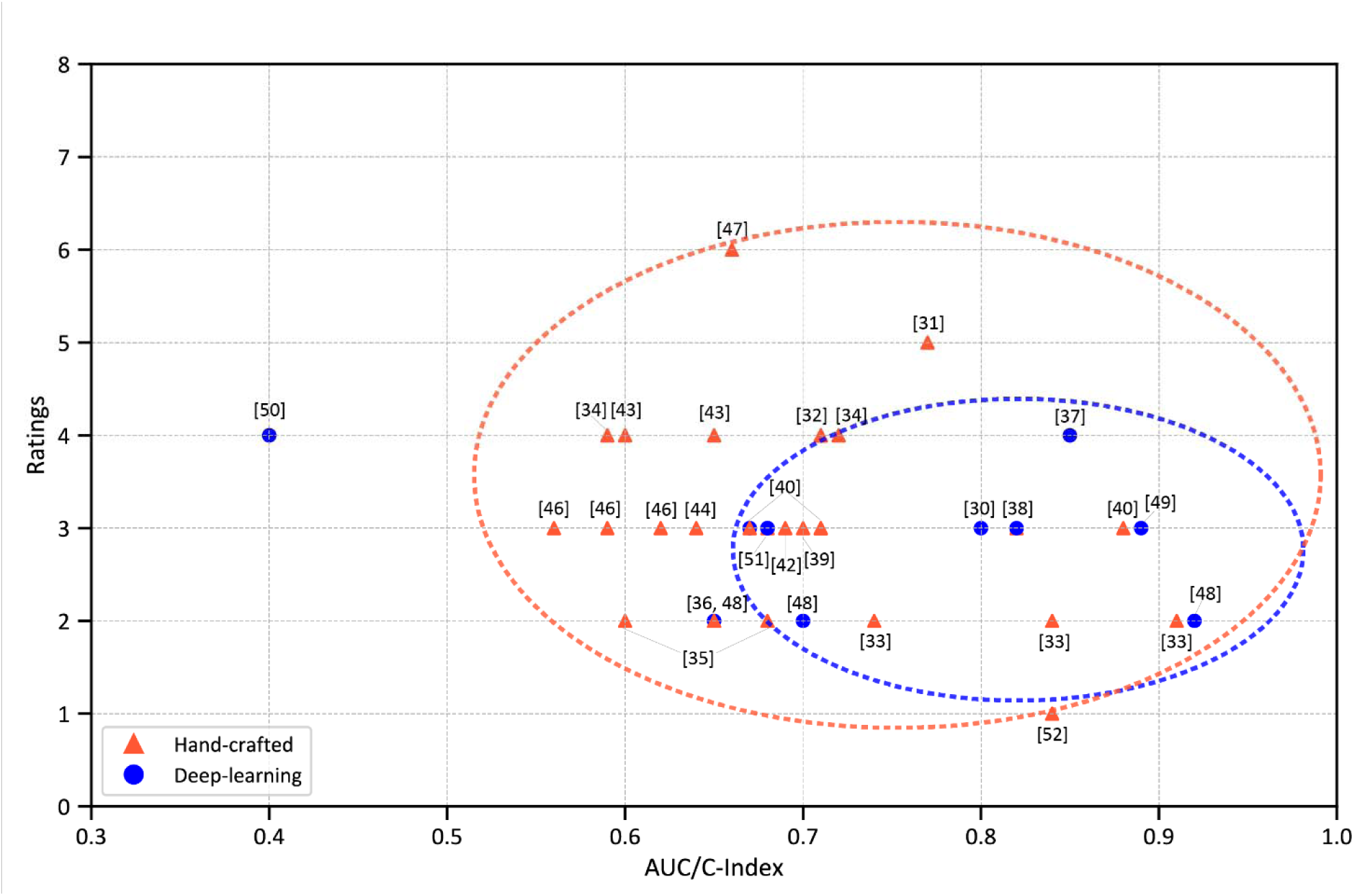
Reported discriminatory metrics (AUC/C-index) of included studies with the number of methodological items rated ‘good’ in this review.

## Discussion

In this systematic review, we summarized the basic characteristics and reported results of studies and rigorously evaluated the methodological quality of studies that included either HC radiomics or DL methods to predict disease outcome in patients treated for head and neck cancer. The models focused on the prediction of recurrent disease and/or survival and were constructed using either HC or DL based radiomics features or both. Only studies that qualified as TRIPOD 3b or higher (had independent validation of the results in an external dataset) were included in the review. While a handful of the studies have reported encouraging results and hinted at their suitability for clinical use, a considerable portion of studies still fall short in their methodological rigor. Future studies can enhance the quality based on the quality checklist provided which allows them to think about employing more robust methodologies and ensuring documentation for wider implementation.

Figure 3 offers an overview of the studies examined in this review, presenting their reported performance metrics alongside ratings from our methodological assessment independently for models validated using either HC or DL based radiomics. The two studies with the highest reported AUC/C index metrics for HC and DL also happen to have very low methodological robustness [33, 48]. Overall, we noted that the DL models exhibited higher performance scores compared to the HC models with the exception of [50] that recorded the lowest performance of AUC/C Index of 0.4. However, despite the higher discriminatory performance, we observed that the methodological aspects were generally better in studies currently using HC radiomics than those involving DL methods. This could be partly attributed to the significant efforts towards standardization of HC radiomics, particularly through initiatives like the Imaging Biomarker Standardization Initiative (IBSI) [53] that has led to clearer definitions, workflows, and best practices. In contrast, deep learning, while rapidly evolving and is sometimes integrated with the HC radiomics workflow, currently lacks the same level of formalized guidelines emphasizing the need for such initiatives.

It would have been optimal if the data collection and statistical analysis protocol for radiomics modeling had been pre-registered. Platforms like ClinicalTrials.gov could serve this purpose, providing transparency in the analysis. Regrettably, none of the studies in this review report such a pre-registration. This may be attributed to the absence of widely available consensus on where such protocols can be registered in advance. We also recommend that, as a radiomics community, we should further promote biomedical modeling registries like the AIMe registry [54]. These platforms should facilitate review, provide suggestions for collaboration, and offer feedback on statistical protocols before initiating a radiomics project study. Transparent registration helps ensure reproducibility and credibility in radiomics research by minimizing biases and clearly defining the methodological framework, including training and validation strategies.

Similarly, we observed that AI prognostication models often overlook the assessments of their clinical implications and applicability for practical use. The evaluation of clinical utility, carried out through methodologies like cost-benefit analysis or decision curve analysis [55, 56], it is imperative for gaining insights on the practical implications of these models. Regrettably, none of the papers included in this review fulfilled both criteria, highlighting a gap in research transparency and pre-analysis protocol documentation.

In our methodological assessment, we evaluated studies based on the clarity of outcome definitions and endpoints. While predicting patient prognosis remains a challenge, to ensure the reliability of the prognostic models, a clear definition of primary endpoint is required that are both valid and reliable. Currently, the endpoints were accepted as defined by the clinicians based on the oncology practices. The endpoints studied include OS, LRR and DM. Most studies lacked clear information for how their clinical endpoints were determined, and whether this was accurately and consistently applied. For instance, when modeling OS, better statistics on the date of death of people that can be prospectively collected is preferred to assess survival interval, as opposed to phone surveys with next-of-kin, but it was not always clear how the important endpoint information was obtained. It could be hospital-based or like in the Netherlands, a national population registry for all births/deaths related information. Overall, there was heterogeneity in the broad definitions of the endpoints and follow-up periods available for survival analysis which could have also contributed to the results varying significantly, with C-Index/AUC values ranging between 0.40 [50] to 0.92 [48]. Notable exceptions to where there was clarity on the clinician-defined endpoints include [31, 32, 47, 50].

Many studies showed [30, 32, 33, 38–40, 43, 47, 49, 51] that a combined model involving clinical factors and imaging features outperforms the results of just the clinical model. These findings suggest that multi-dimensional data possesses greater predictive capability compared to a predictive model constructed solely with mono-dimensional data. However, Le et al. reported that the addition of PET to either the CT or a combination of CT and clinical DL model showed a marked decrease in performance in the models’ predictive capability for all endpoints [51]. It is interesting to note that the same training dataset was used by Goncalves [33] and Vallieres [40].

Clinical research and modelling become highly relevant to the clinician only if the study is accurate and reproducible. The prognostic efficacy of the survival models leveraging radiomics features relies on the utilization of stable and reproducible features, alongside transparent imaging protocols [57, 58]. While studies such as [47] and [37] stand out as exemplary studies with comprehensive and reproducible imaging details, other studies were missing key information such as CT image acquisition and reconstruction parameters.

Most studies also lacked sufficient details for model reproducibility. To reproduce the model, feature engineering and model building are equally important steps. Feature selection methods work by reducing the number of input variables by eliminating redundant features and selecting the most relevant ones for the model. This process significantly enhances model performance, improves interpretability of findings, and addresses generalization issues. In our methodological assessment, we evaluated studies based on their model simplicity and reproducibility. Most HC radiomics studies have clearly outlined their feature reduction and model selection parameters to ensure reproducibility. However, Aerts et al. [39] provided explanations for some statistical methods used but lacked clarity on certain aspects. Zhou et al. (2020) [52] developed multifaceted radiomics models for predicting distant metastases, incorporating both DL and machine learning classifiers. While their feature extraction was conventional, reproducibility steps were not explicitly detailed. For the DL studies, we noted that the model parameters were typically disclosed to ensure reproducibility across most included studies.

It is crucial to understand the biological correlates of features included in the model to improve its interpretability. In some HC radiomics studies we noticed a lack of emphasis on model interpretability, like comparing the model’s performance with established clinical parameters [35, 36, 42, 44, 45]. Goncalves et al. [33] incorporated a combined model of radiomics and clinical parameters, but it should be noted that the clinical parameters were not predictors usually reported in literature for that outcome. While [42, 46] integrate clinical parameters in separate models, a combined model for interpreting the biological implications of the radiomics parameters was absent. In DL, the focus shifts to making the activated regions, from which features influencing the chosen outcome are derived, interpretable for clinicians. Attention maps play a significant role in this context. Except [48], none provided minimal and maximal activation maps. However, it should be emphasized that the activation maps did not correlate the model’s covariates with any known clinical biomarkers. Except [37], other studies trained models incorporating both DL and known clinical features, with a focus to enhance the comprehension of the biological correlates of deep-extracted features.

During our literature review, we encountered papers submitted for the HECKTOR Challenge at MICCAI 2021 and 2022 [59, 60], which focused on automatic head and neck tumor segmentation and outcome prediction in PET/CT images. These papers demonstrated that by integrating radiomics features with machine learning algorithms, valuable insights can be provided into the metabolic and morphological properties of tumors, aiding in the prediction of patient outcomes. The challenge participants were given the same data, and their work centered on applying DL and conventional radiomics to head and neck cancer diagnosis and prognosis, specifically Recurrence-Free Survival (RFS), using FDG-PET/CT images and available clinical data. Despite being highly relevant to our search criteria, these studies were not included as they did not meet the TRIPOD criteria.

We acknowledge several limitations of the current systematic review that future research could address. Firstly, this review was not prospectively registered prior to commencement. Second, we were unable to perform a quantitative meta-analysis owing to the significant heterogeneity in the outcomes analyzed, the methodological and mathematical process involved for HC and DL-based modelling. Instead, we provided a visual synthesis of reported model performance in relation to methodological robustness (Figure 3). Third, despite our rigorous efforts to evaluate methodological procedures using objective criteria, independent raters, and consensus, we believe some degree of subjectivity and potential debatable assessments may remain. Additional detailed notes on methodology are provided in the supplementary table S1 to improve transparency. Lastly, we introduced some inclusion bias by only considering full-text articles in English. This decision was made pragmatically, as all authors of this review are proficient in English, ensuring that the selected material is accessible and understandable to readers who may wish to inspect the individual papers themselves. Additionally, since January 2024 TRIPOD has released an update called TRIPOD+AI. There may be some things in the new TRIPOD that we did not align or incorporate in this present review. However, this may be useful in future work in this question.

## Conclusion

This systematic review provides a critical evaluation of the current state of handcrafted radiomics and deep learning models for prognostication in head and neck squamous cell carcinoma. Despite promising advancements, significant methodological heterogeneity and gaps in reporting standards were identified. The review emphasizes the need for standardized methodologies, including pre-registration of study protocols and detailed reporting of imaging and model development procedures, to improve the reproducibility and clinical utility of these models. Future research should also focus on integrating clinical factors with radiomics features to enhance predictive accuracy and on conducting comprehensive assessments of the clinical implications and cost-effectiveness of these models. Such efforts will be crucial in advancing personalized treatment strategies and improving outcomes for HNSCC patients.

## Data Availability

All data produced in the present work are contained in the manuscript.

## Statements & Declarations

## Funding

Authors acknowledge financial support from the NWO funded personal health Train for RAdiation oncology in India and the Netherlands - TRAIN project (629-002-212), the Stichting Hanarth Fonds and the Proton Therapy Research Infrastructure – ProTRAIT project (RUG 2017-8254) which is supported by the Dutch Cancer Society. Additionally, HMTT acknowledges the DBT/Wellcome Trust India Alliance Early Career Fellowship [Grant number: IA/E/18/1/504306] for the support.

## Competing Interest

Andre Dekker is a founder, shareholder and employee of Medical Data Works B.V. All other authors do not have any competing interest.

## Author Contributions

Varsha Gouthamchand (VG), Louise AF Fonseca (LAAF), Leonard Wee (LW), and Hannah Mary Thomas T (HMTT) contributed to the literature search, study design, eligibility assessment, and manuscript editing. Frank JP Hoebers (FH), as the senior clinician, reviewed the draft to ensure clinical relevance. Rianne Fijten (RF), Andre Dekker (AD), Leonard Wee (LW), and Hannah Mary Thomas T (HMTT) supervised the review process. All authors have read and approved the final version of the manuscript for publication.

## Ethics approval

This review was performed in accordance with the relevant ethical guidelines and regulations of the institution, and did not require any informed consent.

## Consent to publish

Not applicable

