## Supplementary Materials for "Performance of Handcrafted Radiomics versus Deep Learning for Prognosticating Head and Neck Squamous Cell Carcinoma – A Systematic Review with Critical Appraisal of Quantitative Imaging Studies"

**Table S1. Methodological robustness of included studies.**

| Reference | Study pre-registration | Imaging protocol stated in detail | Pre-processing given in detail | Model simplification and reproducibility | Model interpretability | Risk groupings justified | Reliability of model | Exploration of clinical utility |
| --- | --- | --- | --- | --- | --- | --- | --- | --- |
| Aerts:2014 [39] | Poor  Not done | Medium  Imaging protocol not adequate for reproducing the study | Good | Medium  Statistical tests lack enough information for reproducibility of cut-off.  Feature rank stability based on lung datasets. Unsure if they translate to head and neck. | Good | Good | Medium  Outcome was not clearly defined.  Survival data not linked to any reliable follow-up registry  Model Parameters are missing | Poor  Not shown |
| Bogowicz:2020 [45] | Poor  Not done | Medium  Imaging protocol not adequate for reproducing the study | Good | Good | Poor  Not shown | Good | Medium  Outcome was not clearly defined.  Parameters for feature extraction not provided | Poor  Not shown |
| Folkert:2017 [35] | Poor  Not done | Medium  Imaging protocol not adequate for reproducing the study | Medium  Preprocessing details incomplete.  Feature extraction parameters inadequate for reproducibility | Good | Poor  Not shown | Good | Medium  Parameters for feature extraction not provided. | Poor  Not shown |
| Ger:2019 [34] | Poor  Not done | Medium  Imaging protocol not adequate for reproducing the study | Good | Good | Good | Good  . | Medium  Survival data not linked to any reliable follow-up registry  Parameters for feature extraction not provided. | Poor  Not shown |
| Goncalves:2022 [33] | Poor  Not done | Medium  Imaging protocol not adequate for reproducing the study | Good | Good | Medium  Not tested against known clinical predictors | Poor  Not performed | Medium  Outcome was not clearly defined.  Survival data not linked to any reliable follow-up registry  Parameters for feature extraction not provided | Poor  Not shown |
| Keek:2020 [46] | Poor  Not done | Medium  Imaging protocol not adequate for reproducing the study | Good | Good | Medium | Good | Medium  Survival data not linked to any reliable follow-up registry | Poor  Not shown |
| Kim:2022 [31] | Poor  Not done | Medium  Imaging protocol not adequate for reproducing the study | Good | Good | Good | Good | Good | Poor  Not shown |
| Leger:2017 [44] | Poor  Not done | Medium  Imaging protocol not adequate for reproducing the study | Good | Good | Poor  Not shown | Good | Medium  Outcome was not clearly defined.  Survival data not linked to any reliable follow-up registry | Poor  Not shown |
| Leijenaar:2015 [36] | Poor  Not done | Medium  Imaging protocol not adequate for reproducing the study | Poor  Not given | Good  Feature reduction not applicable | Poor  Not shown | Good | Medium  Outcome was not clearly defined.  Survival data not linked to any reliable follow-up registry | Poor  Not shown |
| Lv:2020 [43] | Poor  Not done | Medium  Imaging protocol not adequate for reproducing the study | Good | Good | Good | Good | Medium  Outcome was not clearly defined.  Survival data not linked to any reliable follow-up registry | Poor  Not shown |
| Meneghetti:2021 [47] | Poor  Not done | Good | Good | Good | Good | Good | Good | Poor  Not shown |
| Parmar:2015a [41] | Poor  Not done | Medium  Lung 1, Lung 2, HN1, HN2 similar as Aerts: 2014 | Good | Good | Medium  No combined model. | Good | Medium  Outcome was not clearly defined.  Survival data not linked to any reliable follow-up registry  Parameters for feature extraction not provided | Poor  Not shown |
| Parmar:2015b [42] | Poor  Not done | Medium  Same as Aerts: 2014 | Good  Same as Aerts: 2014 | Good  Same as Aerts: 2014 | Poor  Not shown | Good | Medium  Outcome was not clearly defined.  Survival data not linked to any reliable follow-up registry  Parameters for feature extraction not provided | Poor  Not shown |
| Vallières:2017 [40] | Poor  Not done | Medium  Imaging protocol not adequate for reproducing the study | Poor  Not provided | Good | Good | Good | Medium  Outcome was not clearly defined.  Survival data not linked to any reliable follow-up registry  Cut-off for binary classification of other outcomes was not provided | Poor  Not shown |
| Zhai: 2021 [32] | Poor  Not done | Medium  Imaging protocol not adequate for reproducing the study | Poor  Not provided | Good  Not applicable. | Good | Good  . | Good | Poor  Not shown |
| Zhou:2020 [52] | Poor  Not done | Medium  Same as Vallieres:2017 | Poor  Inadequate for reproducibility | Poor  Feature extraction details for HC features not mentioned | Good | Poor  Not provided | Medium  Outcome was not clearly defined. | Poor  Not shown |
| Cheng:2021 [30] | Poor  Not done | Medium  Imaging protocol not adequate for reproducing the study | Good | Good | Medium  Activation map not available | Good | Medium  Outcome was not clearly defined.  Survival data not linked to any reliable follow-up registry | Poor  Not shown |
| Diamant:2019 [48] | Poor  Not done | Medium  Same as Vallieres:2017 | Good | Good | Medium  Not compared with any known clinical predictors | Poor  Not provided | Medium  Outcome was not clearly defined.  Survival data not linked to any reliable follow-up registry  Cut-off for binary classification of other outcomes was not provided | Poor  Not shown |
| Fujima:2021 [37] | Poor  Not done | Good | Good | Good  . | Medium  Activation map not available | Good | Medium  Incidence rate of outcome not recorded | Poor  Not shown |
| Kazmierski:2023 [38] | Poor  Not done | Medium  Imaging protocol not adequate for reproducing the study | Good  . | Good | Medium  Activation map not available | Good  . | Medium  Outcome was not clearly defined.  Survival data not linked to any reliable follow-up registry | Poor  Not shown |
| Le WT:2022 [51] | Poor  Not done | Medium  Same as Vallieres:2017 | Good | Good | Medium  Activation map not available | Good | Medium  Outcome was not clearly defined.  Survival data not linked to any reliable follow-up registry | Poor  Not shown |
| Lombardo:2021 [49] | Poor  Not done | Medium  Same as Diamant: 2019 | Good | Good | Medium  Activation map not available | Good | Medium  Outcome was not clearly defined. | Poor  Not shown |
| Starke:2020 [50] | Poor  Not done | Medium  Same as Leger:2017 | Good | Good | Medium  Activation map not available | Good | Good | Poor  Not shown |

*Abbreviations: NSCLC - non-small cell lung cancer; HNSCC - head-and-neck squamous cell carcinoma; OPC – Oropharyngeal carcinoma; LPC – laryngeal carcinoma; HPC – hypopharyngeal carcinoma; DL – deep learning; CNN – convolutional neural network; OS – Overall Survival; LRC – Locoregional Control; DM – Distant Metastasis; DFS - Disease-Free Survival; PFS - Progression-Free Survival; LRR – Locoregional Recurrence; LF – Local Failure; DMFS – Distant Metastasis-free Survival; RFS – Recurrence-free Survival; RT – Radiotherapy; CRT – Chemoradiotherapy; IMRT - Intensity-modulated radiation therapy (which also includes volume-modulated arc therapy and helical tomotherapy); CTCAE - Common Terminology Criteria for Adverse Events.*

**(((((((((((((((radiomic*[Text Word]) OR (textur* analys*[Text Word])) OR (feature*[Text Word])) OR (histogram[Text Word])) OR (shape[Text Word])) OR (morpholog*[Text Word])) OR (textur*[Text Word])) OR (image[Text Word])) OR (imaging[Text Word]) AND ((fft[Filter]) AND (humans[Filter]) AND (english[Filter]))) OR ((((((((deep learn*[Text Word]) OR (neural network*[Text Word])) OR (artificial neural[Text Word])) OR (convolutional neural network[Text Word])) OR (CNN[Text Word])) OR (ConvNet[Text Word])) OR (DNN[Text Word])) OR (DLNN[Text Word])) AND ((fft[Filter]) AND (humans[Filter]) AND (english[Filter]))) AND ((((((radiotherapy[Text Word]) OR (radiation therapy[Text Word])) OR (radio therapy[Text Word])) OR (radiation oncology[Text Word])) OR (radiooncology[Text Word])) OR (radio oncology[Text Word]))) AND ((((((PET[Text Word]) OR (positron emission tomography[Text Word])) OR (magnetic resonance[Text Word])) OR (MRI[Text Word])) OR (computed tomography[Text Word])) OR (CT[Text Word]))) AND (head and neck cancer[MeSH Terms])) AND (((((((("Validat"[All Fields] OR (((((((((((((((("predict"[All Fields] OR "predictabilities"[All Fields]) OR "predictability"[All Fields]) OR "predictable"[All Fields]) OR "predictably"[All Fields]) OR "predicted"[All Fields]) OR "predicting"[All Fields]) OR "prediction"[All Fields]) OR "predictions"[All Fields]) OR "predictive"[All Fields]) OR "predictively"[All Fields]) OR "predictiveness"[All Fields]) OR "predictives"[All Fields]) OR "predictivities"[All Fields]) OR "predictivity"[All Fields]) OR "predicts"[All Fields]) AND "ti"[All Fields])) OR "Rule"[All Fields]) OR (((((((((((((((("predict"[All Fields] OR "predictabilities"[All Fields]) OR "predictability"[All Fields]) OR "predictable"[All Fields]) OR "predictably"[All Fields]) OR "predicted"[All Fields]) OR "predicting"[All Fields]) OR "prediction"[All Fields]) OR "predictions"[All Fields]) OR "predictive"[All Fields]) OR "predictively"[All Fields]) OR "predictiveness"[All Fields]) OR "predictives"[All Fields]) OR "predictivities"[All Fields]) OR "predictivity"[All Fields]) OR "predicts"[All Fields]) AND ((("outcome"[All Fields] OR "outcomes"[All Fields]) OR ("risk"[MeSH Terms] OR "risk"[All Fields])) OR ((((((((((((((((("model"[All Fields] OR "model s"[All Fields]) OR "modeled"[All Fields]) OR "modeler"[All Fields]) OR "modeler s"[All Fields]) OR "modelers"[All Fields]) OR "modeling"[All Fields]) OR "modelings"[All Fields]) OR "modelization"[All Fields]) OR "modelizations"[All Fields]) OR "modelize"[All Fields]) OR "modelized"[All Fields]) OR "modelled"[All Fields]) OR "modeller"[All Fields]) OR "modellers"[All Fields]) OR "modelling"[All Fields]) OR "modellings"[All Fields]) OR "models"[All Fields])))) OR (((((((((("history"[MeSH Terms] OR "history"[All Fields]) OR "histories"[All Fields]) OR "history"[MeSH Subheading]) OR ((((("variabilities"[All Fields] OR "variability"[All Fields]) OR "variable"[All Fields]) OR "variable s"[All Fields]) OR "variables"[All Fields]) OR "variably"[All Fields])) OR (((("criteria s"[All Fields] OR "criterias"[All Fields]) OR "standards"[MeSH Subheading]) OR "standards"[All Fields]) OR "criteria"[All Fields])) OR "Scor"[All Fields]) OR ("characteristic"[All Fields] OR "characteristics"[All Fields])) OR (((((((("diagnosis"[MeSH Subheading] OR "diagnosis"[All Fields]) OR "findings"[All Fields]) OR "diagnosis"[MeSH Terms]) OR "finds"[All Fields]) OR "signs and symptoms"[MeSH Terms]) OR ("signs"[All Fields] AND "symptoms"[All Fields])) OR "signs and symptoms"[All Fields]) OR "finding"[All Fields])) OR (("factor"[All Fields] OR "factor s"[All Fields]) OR "factors"[All Fields])) AND ((((((((((((((((((("predict"[All Fields] OR "predictabilities"[All Fields]) OR "predictability"[All Fields]) OR "predictable"[All Fields]) OR "predictably"[All Fields]) OR "predicted"[All Fields]) OR "predicting"[All Fields]) OR "prediction"[All Fields]) OR "predictions"[All Fields]) OR "predictive"[All Fields]) OR "predictively"[All Fields]) OR "predictiveness"[All Fields]) OR "predictives"[All Fields]) OR "predictivities"[All Fields]) OR "predictivity"[All Fields]) OR "predicts"[All Fields]) OR ((((((((((((((((("model"[All Fields] OR "model s"[All Fields]) OR "modeled"[All Fields]) OR "modeler"[All Fields]) OR "modeler s"[All Fields]) OR "modelers"[All Fields]) OR "modeling"[All Fields]) OR "modelings"[All Fields]) OR "modelization"[All Fields]) OR "modelizations"[All Fields]) OR "modelize"[All Fields]) OR "modelized"[All Fields]) OR "modelled"[All Fields]) OR "modeller"[All Fields]) OR "modellers"[All Fields]) OR "modelling"[All Fields]) OR "modellings"[All Fields]) OR "models"[All Fields])) OR (((("decision"[All Fields] OR "decision s"[All Fields]) OR "decisions"[All Fields]) OR "decisive"[All Fields]) OR "decisively"[All Fields])) OR "Identif"[All Fields]) OR "Prognos"[All Fields]))) OR ((((("decision"[All Fields] OR "decision s"[All Fields]) OR "decisions"[All Fields]) OR "decisive"[All Fields]) OR "decisively"[All Fields]) AND ((((((((((((((((((("model"[All Fields] OR "model s"[All Fields]) OR "modeled"[All Fields]) OR "modeler"[All Fields]) OR "modeler s"[All Fields]) OR "modelers"[All Fields]) OR "modeling"[All Fields]) OR "modelings"[All Fields]) OR "modelization"[All Fields]) OR "modelizations"[All Fields]) OR "modelize"[All Fields]) OR "modelized"[All Fields]) OR "modelled"[All Fields]) OR "modeller"[All Fields]) OR "modellers"[All Fields]) OR "modelling"[All Fields]) OR "modellings"[All Fields]) OR "models"[All Fields]) OR (((((((("ambulatory care facilities"[MeSH Terms] OR (("ambulatory"[All Fields] AND "care"[All Fields]) AND "facilities"[All Fields])) OR "ambulatory care facilities"[All Fields]) OR "clinic"[All Fields]) OR "clinic s"[All Fields]) OR "clinical"[All Fields]) OR "clinically"[All Fields]) OR "clinicals"[All Fields]) OR "clinics"[All Fields])) OR (("logistic models"[MeSH Terms] OR ("logistic"[All Fields] AND "models"[All Fields])) OR "logistic models"[All Fields])))) OR (((((((((((("prognostic"[All Fields] OR "prognostical"[All Fields]) OR "prognostically"[All Fields]) OR "prognosticate"[All Fields]) OR "prognosticated"[All Fields]) OR "prognosticates"[All Fields]) OR "prognosticating"[All Fields]) OR "prognostication"[All Fields]) OR "prognostications"[All Fields]) OR "prognosticator"[All Fields]) OR "prognosticators"[All Fields]) OR "prognostics"[All Fields]) AND (((((((((("history"[MeSH Terms] OR "history"[All Fields]) OR "histories"[All Fields]) OR "history"[MeSH Subheading]) OR ((((("variabilities"[All Fields] OR "variability"[All Fields]) OR "variable"[All Fields]) OR "variable s"[All Fields]) OR "variables"[All Fields]) OR "variably"[All Fields])) OR (((("criteria s"[All Fields] OR "criterias"[All Fields]) OR "standards"[MeSH Subheading]) OR "standards"[All Fields]) OR "criteria"[All Fields])) OR "Scor"[All Fields]) OR ("characteristic"[All Fields] OR "characteristics"[All Fields])) OR (((((((("diagnosis"[MeSH Subheading] OR "diagnosis"[All Fields]) OR "findings"[All Fields]) OR "diagnosis"[MeSH Terms]) OR "finds"[All Fields]) OR "signs and symptoms"[MeSH Terms]) OR ("signs"[All Fields] AND "symptoms"[All Fields])) OR "signs and symptoms"[All Fields]) OR "finding"[All Fields])) OR (("factor"[All Fields] OR "factor s"[All Fields]) OR "factors"[All Fields])) OR ((((((((((((((((("model"[All Fields] OR "model s"[All Fields]) OR "modeled"[All Fields]) OR "modeler"[All Fields]) OR "modeler s"[All Fields]) OR "modelers"[All Fields]) OR "modeling"[All Fields]) OR "modelings"[All Fields]) OR "modelization"[All Fields]) OR "modelizations"[All Fields]) OR "modelize"[All Fields]) OR "modelized"[All Fields]) OR "modelled"[All Fields]) OR "modeller"[All Fields]) OR "modellers"[All Fields]) OR "modelling"[All Fields]) OR "modellings"[All Fields]) OR "models"[All Fields])))) OR (((((((((((("stratification"[All Fields] OR "stratifications"[All Fields]) OR (("roc curve"[MeSH Terms] OR ("roc"[All Fields] AND "curve"[All Fields])) OR "roc curve"[All Fields])) OR ((((((((((((((((((("discriminabilities"[All Fields] OR "discriminability"[All Fields]) OR "discriminable"[All Fields]) OR "discriminably"[All Fields]) OR "discriminance"[All Fields]) OR "discriminant"[All Fields]) OR "discriminants"[All Fields]) OR "discriminate"[All Fields]) OR "discriminated"[All Fields]) OR "discriminates"[All Fields]) OR "discriminating"[All Fields]) OR "discrimination, psychological"[MeSH Terms]) OR ("discrimination"[All Fields] AND "psychological"[All Fields])) OR "psychological discrimination"[All Fields]) OR "discrimination"[All Fields]) OR "discriminations"[All Fields]) OR "discriminative"[All Fields]) OR "discriminatively"[All Fields]) OR "discriminator"[All Fields]) OR "discriminators"[All Fields])) OR ((((((((((((((((((("discriminabilities"[All Fields] OR "discriminability"[All Fields]) OR "discriminable"[All Fields]) OR "discriminably"[All Fields]) OR "discriminance"[All Fields]) OR "discriminant"[All Fields]) OR "discriminants"[All Fields]) OR "discriminate"[All Fields]) OR "discriminated"[All Fields]) OR "discriminates"[All Fields]) OR "discriminating"[All Fields]) OR "discrimination, psychological"[MeSH Terms]) OR ("discrimination"[All Fields] AND "psychological"[All Fields])) OR "psychological discrimination"[All Fields]) OR "discrimination"[All Fields]) OR "discriminations"[All Fields]) OR "discriminative"[All Fields]) OR "discriminatively"[All Fields]) OR "discriminator"[All Fields]) OR "discriminators"[All Fields])) OR ("c"[All Fields] AND (((("statistic"[All Fields] OR "statistic s"[All Fields]) OR "statistical"[All Fields]) OR "statistically"[All Fields]) OR "statistics"[All Fields]))) OR ("c"[All Fields] AND (((("statistic"[All Fields] OR "statistic s"[All Fields]) OR "statistical"[All Fields]) OR "statistically"[All Fields]) OR "statistics"[All Fields]))) OR (((("area under curve"[MeSH Terms] OR (("area"[All Fields] AND "under"[All Fields]) AND "curve"[All Fields])) OR "area under curve"[All Fields]) OR (("area"[All Fields] AND "under"[All Fields]) AND "curve"[All Fields])) OR "area under the curve"[All Fields])) OR "auc"[All Fields]) OR (((((((((("calibrant"[All Fields] OR "calibrants"[All Fields]) OR "calibrate"[All Fields]) OR "calibrated"[All Fields]) OR "calibrates"[All Fields]) OR "calibrating"[All Fields]) OR "calibration"[MeSH Terms]) OR "calibration"[All Fields]) OR "calibrations"[All Fields]) OR "calibrator"[All Fields]) OR "calibrators"[All Fields])) OR ((((((((((((("indicate"[All Fields] OR "indicated"[All Fields]) OR "indicates"[All Fields]) OR "indicating"[All Fields]) OR "indicative"[All Fields]) OR "indicatives"[All Fields]) OR "indicators and reagents"[Pharmacological Action]) OR "indicators and reagents"[MeSH Terms]) OR ("indicators"[All Fields] AND "reagents"[All Fields])) OR "indicators and reagents"[All Fields]) OR "indicator"[All Fields]) OR "indicators"[All Fields]) OR "indice"[All Fields]) OR "indices"[All Fields])) OR ((((((("algorithm s"[All Fields] OR "algorithmic"[All Fields]) OR "algorithmically"[All Fields]) OR "algorithmics"[All Fields]) OR "algorithmization"[All Fields]) OR "algorithms"[MeSH Terms]) OR "algorithms"[All Fields]) OR "algorithm"[All Fields])) OR ((((((((("multivariable"[All Fields] OR "multivariables"[All Fields]) OR "multivariably"[All Fields]) OR "multivariance"[All Fields]) OR "multivariant"[All Fields]) OR "multivariate"[All Fields]) OR "multivariated"[All Fields]) OR "multivariately"[All Fields]) OR "multivariates"[All Fields]) OR "multivariative"[All Fields]))) OR (((("predict*"[Title/Abstract] OR "predictive value of tests"[MeSH Terms]) OR "scor*"[Title/Abstract]) OR "observ*"[Title/Abstract]) OR "observer variation"[MeSH Terms])) AND ((fft[Filter]) AND (humans[Filter]) AND (english[Filter]) AND (2010:2020[pdat]))) NOT (review[Publication Type]) AND ((fft[Filter]) AND (humans[Filter]) AND (english[Filter]) AND (2010:2020[pdat]))) NOT ((review[Title]) OR (review[Publication Type]) AND ((fft[Filter]) AND (humans[Filter]) AND (english[Filter])))**

**Text Box 1.** Pubmed Search String
